# Risk factors for clinical progression in patients with COVID-19: a retrospective study of electronic health record data in the United Kingdom

**DOI:** 10.1101/2020.05.11.20093096

**Authors:** Robert A. Fletcher, Thomas Matcham, Marta Tibúrcio, Arseni Anisimovich, Stojan Jovanović, Luca Albergante, Nadia Lipunova, Anne Hancock, Lucy Mackillop, Lionel Tarassenko, Alex McCarthy, Marcela P. Vizcaychipi, Rabia Tahir Khan

## Abstract

**Background:** The novel coronavirus disease 2019 (COVID-19) outbreak presents a significant threat to global health. A better understanding of patient clinical profiles is essential to drive efficient and timely health service strategies. In this study, we aimed to identify risk factors for a higher susceptibility to symptomatic presentation with COVID-19 and a transition to severe disease.

**Methods:** We analysed data on 2756 patients admitted to Chelsea & Westminster Hospital NHS Foundation Trust between 1^st^ January and 23rd April 2020. We compared differences in characteristics between patients designated positive for COVID-19 and patients designated negative on hospitalisation and derived a multivariable logistic regression model to identify risk factors for predicting risk of symptomatic COVID-19. For patients with COVID-19, we used univariable and multivariable logistic regression to identify risk factors associated with progression to severe disease defined by: 1) admission to the hospital’s AICU, 2) the need for mechanical ventilation, 3) in-hospital mortality, and 4) at least one measurement of elevated D-dimer (≥1,000 μg/L) indicative of increased risk of venous thromboembolism.

**Results:** The patient population consisted of 1148 COVID-19 positive and 1608 COVID-19 negative patients. Age, sex, self-reported ethnicity, C-reactive protein, white blood cell count, respiratory rate, body temperature, and systolic blood pressure formed the most parsimonious model for predicting risk of symptomatic COVID-19 at hospital admission. Among 1148 patients with COVID-19, 116 (10.1%) were admitted to the AICU, 71 (6.2%) required mechanical ventilation, 368 (32.1%) had at least one record of D-dimer levels ≥1,000 μg/L, and 118 patients died. In the multivariable logistic regression, age (OR = 0.953 per 1 year, 95% CI: 0.937-0.968) C-reactive protein (OR = 1.004 per 1 mg/L, 95% CI: 1.002-1.007), and white blood cell counts (OR = 1.059 per 10^9^/L, 95% CI: 1.010-1.111) were found to be associated with admission to the AICU. Age (OR = 0.973 per 1 year, 95% CI: 0.955-0.990), C-reactive protein (OR = 1.003 per 1 mg/L, 95% CI: 1.000-1.006) and sodium (OR = 0.915 per 1 mmol/L, 0.868-0.962) were associated with mechanical ventilation. Age (OR = 1.023 per 1 year, 95% CI: 1.004-1.043), CRP (OR = 1.004 per 1 mg/L, 95% CI: 1.002-1.006), and body temperature (OR = 0.723 per 1°C, 95% CI: 0.541-0.958) were associated with elevated D-dimer. For mortality, we observed associations with age (OR = 1.060 per 1 year, 95% CI: 1.040-1.082), female sex (OR = 0.442, 95% CI: 0.442, 95% CI: 0.245-0.777), Asian ethnic background (OR = 2.237 vs White ethnic background, 95% CI: 1.111-4.510), C-reactive protein (OR = 1.004 per 1 mg/L, 95% CI: 1.001-1.006), sodium (OR = 1.038 per 1 mmol/L, 95% CI: 1.001-1.006), and respiratory rate (OR = 1.054 per 1 breath/min, 95% CI: 1.024-1.087).

**Conclusion:** Our analysis suggests there are several demographic, clinical and laboratory findings associated with a symptomatic presentation of COVID-19. Moreover, significant associations between patient deterioration were found with age, sex and specific blood markers, chiefly C-reactive protein, and could help early identification of patients at risk of poorer prognosis. Further work is required to clarify the extent to which our observations are relevant beyond current settings.

## Introduction

The World Health Organization has classified the coronavirus disease 2019 (COVID-19) pandemic as a public health emergency of international concern. However, many questions regarding epidemiological patterns and spectrum of the disease are yet to be answered. It is estimated that approximately 81% of all COVID-19 patients develop uncomplicated or mild disease, while 14% develop severe illness, and 5% require treatment in a hospital adult intensive care unit (AICU) (1, 2). Risk factors for inferior prognoses are critical to identify, in order to facilitate informed decision-making on prevention measures and patient care by policy makers, epidemiologists, and clinicians.

Recently published observational studies have reported associations between increased mortality in COVID-19 patients with older age, male sex, and history of certain comorbidities, including cardiovascular disease, chronic respiratory diseases, hypertension, diabetes, and obesity (2-5). Further, several biomarkers measured at the time of hospitalisation have been linked with greater disease severity. These include C-reactive protein (CRP), creatinine, creatine kinase, lactate dehydrogenase (LDH), troponin I, N-terminal pro-brain natriuretic peptide (NT-pro BNP), liver function biomarkers alanine aminotransferase (ALT) and aspartate transferase (AST), and numbers of lymphocytes circulating in the blood (3, 6, 7). While these studies provide an important overview of the characteristics of COVID-19 patients, most comprise relatively small sample sizes and are therefore likely to be hindered by low statistical power. Further limitations include systemic underreporting of prior disease history by study participants, as well as insufficient adjustment for confounders in statistical analyses (8). Moreover, most of these studies were conducted in countries that were first afflicted by COVID-19, principally China (2-5, 8-10). Data from other populations is still therefore limited, and further research is required to corroborate all findings to better to characterise the clinical profile of patients with COVID-19.

In this study, we report a retrospective analysis of electronic health record (EHR) data for patients in the United Kingdom who were admitted to the emergency department of a National Health Service (NHS) Trust. These data provided key information on patients’ clinical profile at the time of COVID-19 diagnosis, as well as on historic data for comorbidities and in-hospital prescribed medications. We aimed to compare clinical characteristics at the time of hospitalization between patients with confirmed COVID-19 and patients in whom COVID-19 was not detected, as well as to investigate whether any clinical risk factors are associated with inferior prognoses among patients with COVID-19.

## Methods

### Study population and data collection

For this retrospective observational study, we obtained anonymised EHR data for patients admitted to the emergency department of Chelsea & Westminster Hospital NHS Foundation Trust between 1^st^ January 2020 and 23^rd^ April 2020. The EHR data collected during patients’ hospitalisation period included biochemistry test results, vital sign measurements, and data recorded in the AICU for any patients with severe disease manifestations admitted to the unit for more aggressive therapy (Figure 1). Information on previously recorded diagnoses and in-hospital administered medication was also available. Prior diagnoses were coded in accordance with the International Classification of Diseases, tenth revision (ICD-10). All analyses were restricted to patients aged 16 years or older at the time of their most recent hospital admission (assumed to be the ‘target’ admission at which there were concerns of COVID-19). Patients with incomplete or missing data for risk factors of interest identified through a review of published literature (Supplementary Table 1) were excluded from baseline comparisons and regression analyses.

**Figure 1.**
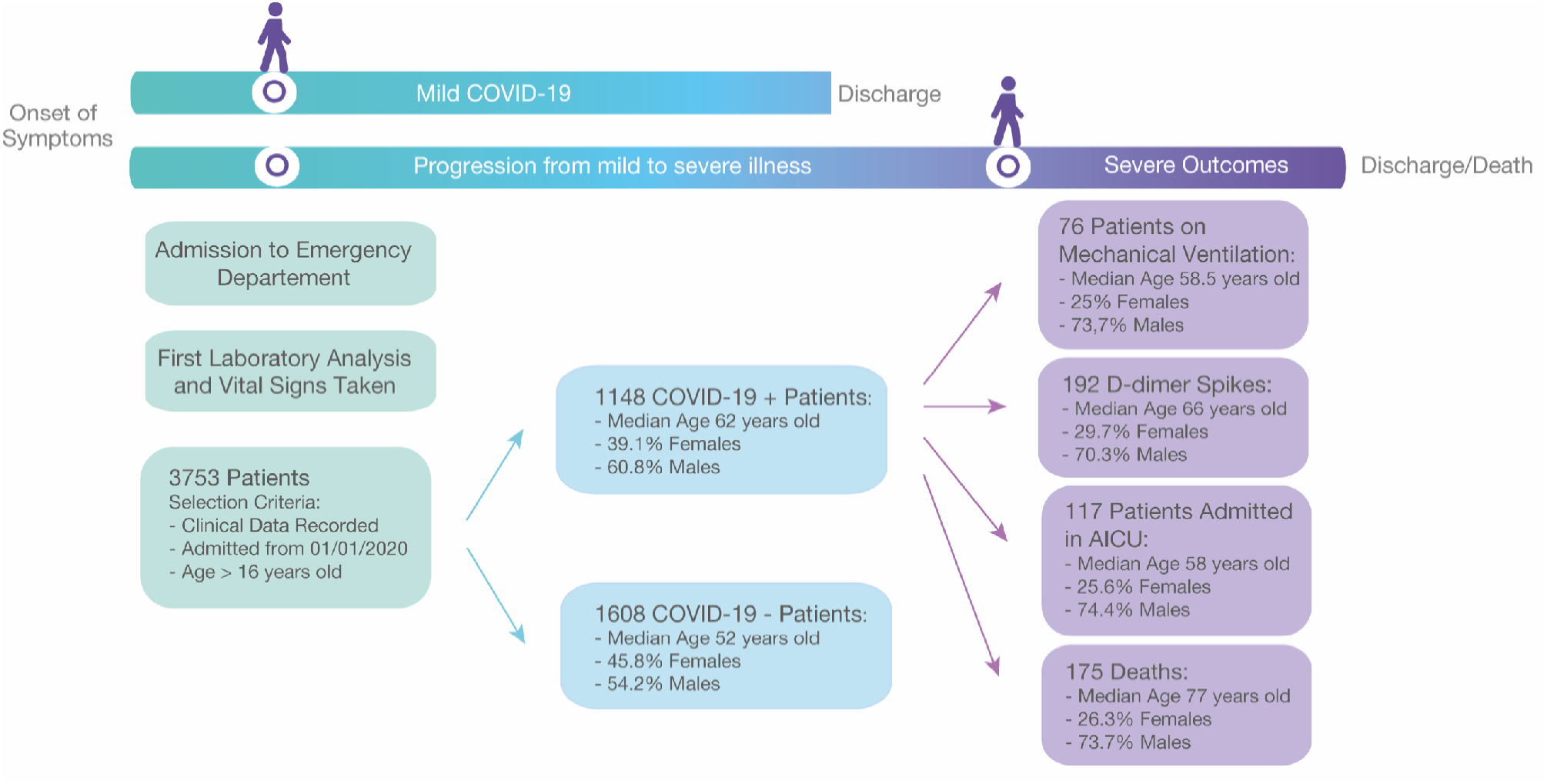
Illustration of the patient pathway, since hospital admission, and analysed patient cohorts. Schematic of the patient pathway after admission in the hospital emergency department, as well as of the number of patients used for the purpose of the retrospective analysis. The initial 3753 patients were classified as COVID-19 positive or negative and the clinical information of the COVID-19 positive patients was used to analyse four severe clinical outcomes (admission to AICU, mechanical ventilation and D-dimer ≥ 1000μg/L, and in-hospital mortality).

### COVID-19 status definition and studied outcomes

Patients were classified as “positive” or “negative” for COVID-19 at the time of admission to the hospital emergency department (Figure 1) following a laboratory confirmation of SARS-CoV-2 by quantitative RT-PCR (qRT-PCR). Patients were classified as “suspected” if there were awaiting a test result or had received a negative test result but exhibited symptoms indicative of COVID-19 during clinical triage. To evaluate risk factors associated with clinical deterioration in patients with COVID-19, four endpoints were defined in this study; specifically: 1) a recorded admission to the hospital AICU, 2) a recorded use of mechanical ventilation for respiratory assistance (defined as administration of one of the following oxygen devices: ‘bipap’, ‘heliox normal’, ‘ventilator’, ‘tracheostomy’, or ‘t-piece’), 3) in-hospital mortality and 4), at least one D-dimer measurement ≥ 1,000 μg/L recorded during the patient’s hospital admission. This threshold for D-dimer was selected as it is linked to greater risk of venous thromboembolism (VTE)(11).

### Statistical analysis

To identify potential differences between COVID-19 positive and negative patients, we compared demographic characteristics (age, sex, and self-reported ethnicity), prior history of medical diagnoses recorded in the Trust within the past 16 years (from March 2004 to present), prior history of treatments administered in the Trust within the past 10 years (from August 2008 to present), and first available laboratory measurements and vital sign observations recorded during patients’ most recent hospital admission.

To assess statistically significant differences in these characteristics, we used Chi-squared tests for comparison of categorical variables, Student’s t-tests for normally distributed continuous variables, and Wilcoxon rank-sum tests for non-normally distributed continuous variables. We also computed standardised mean differences (SMDs) for log-transformed laboratory measurements and vital sign observations to quantify the magnitude of difference in these characteristics between patient groups. When describing the distribution of categorical variables, number and percentage of patients in each category were reported. For continuous variables, the sample mean and standard deviation (SD), or the sample median and interquartile range (IQR) were reported as appropriate.

We further developed a multivariable logistic regression model to identify prognostic factors that can be used to predict risk of COVID-19 in patients admitted to the hospital. This model was developed in patients with a confirmed COVID-19 status (either positive or negative) at the time of admission to the hospital. Variables included in the model were selected by backwards elimination using the Bayesian Information Criterion (12, 13), a likelihood measure which penalises model complexity to prevent overfitting. We then conducted internal validation of the model with both 10-fold cross validation in the sample of patients in which the model was developed, as well as with an internal-external validation approach in those patients for whom COVID-19 status had not initially been confirmed (coded as “suspected” in earlier revisions of the data) but then transitioned to known COVID-19 status at a later stage in their hospital admission period following receipt of a valid test result. We calculated discrimination performance of the model by computing the area under the receiver operating characteristics curve (AUC), which measures the ability of the model to differentiate between those at higher and lower risk of disease status.

In patients positive for COVID-19, we used univariable and multivariable logistic regression to explore the association of measured risk factors, principally age, sex, self-reported ethnicity, laboratory measurements, and vital sign observations, with transfer to the hospital’s AICU, the need for mechanical ventilation, a recorded D-dimer measurement ≥1,000 μg/L, and in-hospital mortality, separately. To avoid the risk of overfitting, we limited the number of variables included in each model to maintain an events-per-variable (EPV) ratio of >10 (14). Fisher’s exact test was also used to compute odds ratio (OR) associations of D-dimer levels <1000 μg/L and ≥1,000 μg/L with AICU admission, mechanical ventilation and mortality. ORs were considered statistically significant at a threshold of p<0.05 by Wald test. Multiple hypothesis testing was adjusted by applying the Holm-Bonferroni correction method.

The inclusion of interaction terms with age and sex was assessed by quantifying their statistical significance within all models at a threshold of p<0.05 by likelihood ratio test, and the linearity of continuous variables was assessed by visual inspection of ordered categorical partial effects plots. All analyses were performed with R version 3.6.0, two-sided p values, and 95% confidence intervals (CIs).

## Results

### Descriptive patient characteristics

A total of 3753 patients suspected for COVID-19 were admitted to the emergency department of Chelsea & Westminster Hospital NHS Foundation Trust (Figure 2). Of the total population, 3663 patients had laboratory measurements and vital sign observations recorded during their admission. Following exclusion of patients younger than 16 years, the final sample comprised 1148 patients identified as positive for COVID-19 and 1608 identified as negative (Figure 1).

**Figure 2.**
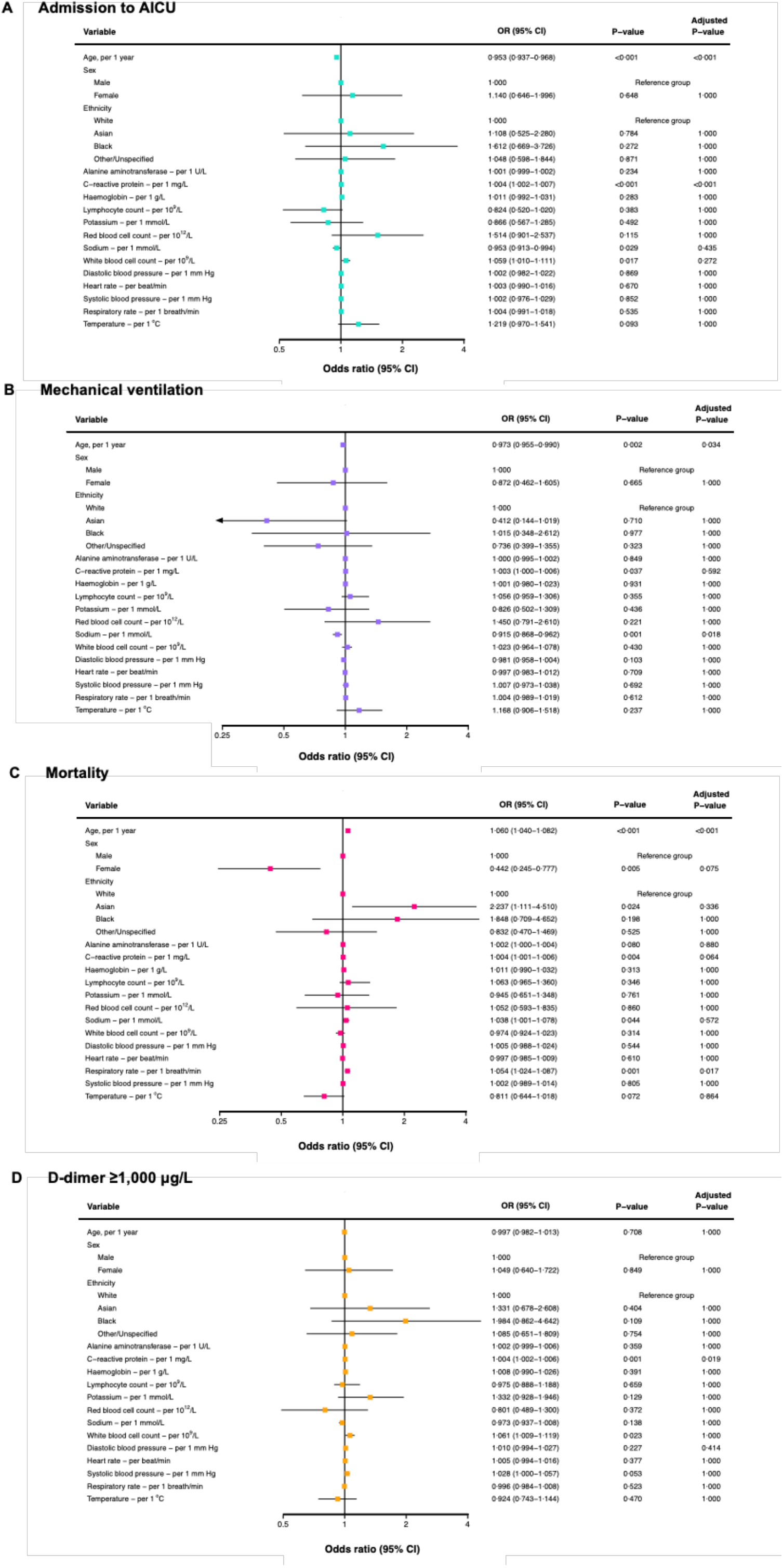
Multivariable-adjusted associations of risk factors with admission to the hospital’s AICU, administration of mechanical ventilation, in-hospital mortality, and D-dimer ≥1000μg/L. Data are odds ratio (95% CI). Adjusted P-values denote those corrected using the Holm-Bonferroni method for multiple testing.

Selected baseline characteristics for the patient population are presented in Supplementary Table 1. On average, patients designated positive for COVID-19 were older (mean age (SD), 61.2 years (20.1) vs 51.4 years (21.9); p<0.001) and are likely to be male (60.8% vs 45.8%; p<0.001) when compared with patients designated negative for COVID-19. Although the population was predominantly of White or unspecified ethnic background, there was a higher proportion of patients with self-reported Asian (14.1% vs 11.3%) and Black (6.1% vs 3.6%) ethnic backgrounds among those patients identified as positive for COVID-19 (Supplementary Table 1).

Information on medical history attributed in the Trust was available for 548 (41.7%) COVID-19 positive and 583 (58.4%) COVID-19 negative patients. Patients with COVID-19 were more likely to have been previously diagnosed with hypertension (50.4% vs 44.1%; p=0.001) and type-II diabetes (28.3% vs 22.1%; p=0.021) (Supplementary Table 1). No significant differences were observed for history of previously administered medication classes (as defined in the British National Formulary (BNF) pharmaceutical reference book for medications available on the UK NHS) (Supplementary Table 1).

Multiple laboratory measurements obtained at the time of hospitalisation showed statistically significant differences between COVID-19 positive and COVID-19 negative patient groups (Supplementary Table 1). In patients identified as positive for COVID-19, we observed higher serum levels of CRP (median [IQR], 101.0 [41.6, 176.8] vs 44.9 [12.8, 114.8] mg/L; p<0.001), LDH (median [IQR], 405 [280, 571] vs 296 [213, 372] IU/L; p<0.001), creatinine (median [IQR], 87.0 [68.0, 118.0] vs 76.0 [62.0, 111.0] µmol/Litre; p<0.001), creatine kinase (median [IQR], 145.5 [66.0, 357.0] vs 89.0 [50.0, 196.5] IU/L; p<0.001), and ferritin (median [IQR], 542.0 [242.0, 1143.0] vs 220.5 [86.5, 520.5] ng/mL; p<0.001).

For key liver function biomarkers, higher serum levels of alanine aminotransferase (ALT; median [IQR], 30.0 U/L [18.0, 48.8] vs 22.0 U/L [14.0, 40.0]; p<0.001) and lower levels of alkaline phosphatase (ALP; median [IQR], 83.0 [64.0, 113.0] vs 91.0 [68.0, 121.0] U/L; p<0.001) were observed in patients with COVID-19.

Absolute numbers of key immune cells were systemically lower in patients with COVID-19, in particular lymphocytes (median [IQR], 0.9 [0.6, 1.4] vs 1.3 [0.8, 1.9] x10^9^/L; p<0.001), neutrophils (median [IQR], 5.7 [4.0, 8.1] vs 6.9 [4.8, 10.4] x10^9^/L; p<0.001), and monocytes (median [IQR], 0.5 [0.4, 0.7] vs 0.7 [0.5, 1.0] x10^9^/L; P<0.001).

For biomarkers of blood coagulation in patients with COVID-19, we observed reduced absolute numbers of platelets (median [IQR], 222 [169, 288] vs 251 (193, 330) x10^9^/L; p<0.001), and concurrent increases in activated partial thromboplastin time (median [IQR], 32.0 [29.2, 35.6] vs 30.5 [27.9, 34.4] seconds; p<0.001) and mean platelet volume (median [IQR], 215.00 [167.00, 280.00] vs 8.30 [7.50, 9.00] fL; p<0.001) (Supplementary Table 1).

Further, significant differences between COVID-19 positive and negative patients were observed for most vital signs (Supplementary Table 1). Respiratory rate (median [IQR] 23.0 [19.0, 28.0] vs 20.0 [18.0, 24.0] breaths/min; p<0.001), total national early warning score version 2 (NEWS-2; median [IQR], 5.0 [2.0, 7.0] vs 3.0 [1.0, 5.0]; p<0.001), body temperature (median [IQR], 37.2 [36.4, 37.8] vs 36.6 [36.1, 37.3] °C; p<0.001) were all higher in patients designated positive for COVID-19. Among patients with recorded use of a mechanical ventilator for respiratory assistance, oxygen flow rate (median [IQR], 3.0 [0.0, 15.0] vs 1.0 [0.0, 4.0] L/min; p<0.001) was higher in patients with COVID-19.

### Association of risk factors with symptomatic COVID-19

The logistic regression model to identify prognostic factors with potential use for predicting risk of COVID-19 in patients admitted to the hospital was developed in 806 patients identified as COVID-19 positive and 768 patients identified as negative at admission. Following variable selection via the Bayesian Information Criterion (BIC), the most parsimonious model included age, sex, self-reported ethnicity, two blood biomarkers (CRP and white blood cell numbers), and three vital sign measurements (respiratory rate, body temperature, and systolic blood pressure) (Table 1). To facilitate independent external validation of the logistic regression model, multivariable-adjusted regression coefficients, ORs, and model intercept are presented in Table 1. In 10-fold cross validation, the optimism-adjusted AUC of the model was computed at 0.76 (95% CI: 0.71-0.81). In the internal-external validation data set of 143 COVID-19 suspected patients for whom COVID-19 status was not initially known but ascertained later, model performance was only marginally reduced, with an AUC of 0.74 (95% CI: 0.70-0.77).

**Table 1.**
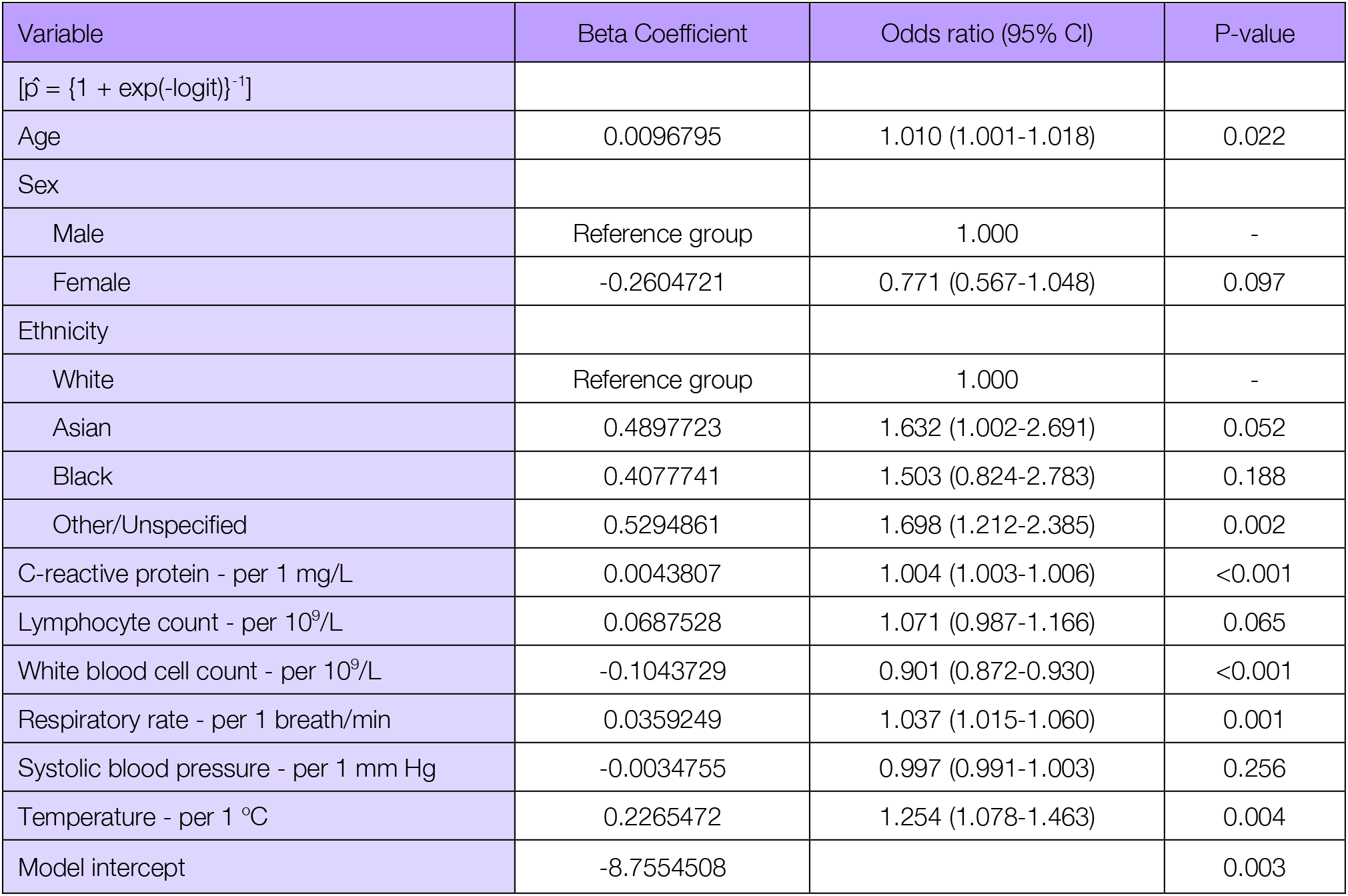
Beta coefficients and odds ratios for the multivariable logistic regression model to predict risk of presentation with symptomatic COVID-19.

## Univariable and multivariable logistic regression

### Admission to AICU

In univariable analyses minimally adjusted for age and sex, we found six blood biomarkers and two vital sign measurements significantly associated with patient admission to the hospital’s AICU (Supplementary Table 2). Among biomarkers, the odds of admission to AICU were higher in those with higher serum concentrations of CRP (OR = 1.006 per 1 mg/L, 95% CI: 1.004-1.008), higher haemoglobin (OR = 1.017 per 1 g/L, 95% CI: 1.006-1.029), greater absolute numbers of red blood cells (RBCs; OR = 1.696 per 10^12^/L, 95% CI: 1.249-2.322), and greater absolute numbers of white blood cells (OR = 1.044 per 10^9^/L, 95% CI: 1.009-1.080). Lower odds of admission to the AICU was observed for higher serum levels of sodium (OR = 0.949 per 1 mmol/L, 95% CI: 0.911-0.987). For vital signs, both higher body temperature (OR = 1.303 per 1°C, 95% CI: 1.082-1.595) and higher respiratory rate (OR = 1.025 per 1 breath/min, 95% CI: 1.001-1.049) were associated with increased odds of AICU admission.

A total of 628 patients with complete data and 112 cases of admission to the AICU, were used to develop the multivariable logistic regression model. Following multivariable-adjustment, most associations were attenuated, with only increases in serum levels of CRP (OR = 1.004 per 1 mg/L, 95% CI: 1.002-1.007) and numbers of white blood cells (OR = 1.059 per 10^9^/L, 95% CI: 1.010-1.111) associated with greater odds of admission to the AICU (Figure 2). Conversely, older age (OR = 0.953 per 1 year, 95% CI: 0.937-0.968) and increased serum levels of sodium (OR = 0.953 per 1 mmol/L, 95% CI: 0.913-0.994) were associated with lower odds of admission to the AICU. No significant associations with sex or ethnicity were observed. Following adjustment of P-values in the multivariable model with the Holm-Bonferroni method, associations with age and CRP levels remained statistically significant (Figure 2).

### Mechanical ventilation

In the univariable analysis, we observed increased odds of mechanical ventilation in those with higher levels of CRP (OR = 1.004 per 1 mg/L, 95% CI: 1.001-1.049) and lower odds with higher serum levels of sodium (OR = 0.923 per 1 mmol/L, 95% CI: 0.879-0.966) (Supplementary Table 2).

For the multivariable logistic regression model, 663 patients with 70 cases of mechanical ventilation and complete data for all risk factors of interest were included. On mutual adjustment in the multivariable model, our results showed that associations of mechanical ventilation with both CRP (OR = 1.003 per 1 mg/L, 95% CI: 1.000-1.006) and sodium levels (OR = 0.915 per 1 mmol/L, 0.868-0.962) remained significant (Figure 2). Older age was also associated with decreased odds of mechanical ventilation (OR = 0.973 per 1 year, 95% CI: 0.955-0.990). No significant associations were observed with sex or ethnicity. Associations with both age and sodium levels remained statistically significant following adjustment of P-values with the Holm-Bonferroni method (Figure 2).

### Mortality

In age- and sex-adjusted univariable analyses, higher odds of mortality were observed in patients with increased CRP levels (OR = 1.003 per 1 mg/L, 95% CI: 1.001-1.006), increased sodium levels (OR = 1.036 per 1 mmol/L, 95% CI: 1.004-1.071), and higher respiratory rate (OR = 1.061 per 1 breath/min, 95% CI: 1.035-1.090) (Supplementary Table 2).. Relative to patients of White ethnic background, we also observed approximately 2-fold greater odds of mortality in patients of Black (OR = 2.205, 95% CI: 0.905-5.199) and Asian (OR = 2.354, 95% CI: 1.232-4.503) ethnic backgrounds, however only the association with Asian patients was statistically significant (Supplementary Table 2).

In 493 patients with 118 cases of mortality, similar associations were observed following multivariable adjustment. Patients with older age (OR = 1.060 per 1 year, 95% CI: 1.040-1.082) and Asian ethnic background (OR = 2.237, 95% CI: 1.111-4.510) had higher odds of mortality, as did patients with higher CRP (OR = 1.004 per 1 mg/L, 95% CI: 1.001-1.006), sodium (OR = 1.038 per 1 mmol/L, 95% CI: 1.001-1.006), and respiratory rate. (OR = 1.054 per 1 breath/min, 95% CI: 1.024-1.087). Female patients were at over 50% lower odds of mortality relative to male patients (OR = 0.442, 95% CI: 0.442, 95% CI: 0.245-0.777). Following the adjustment for multiple hypothesis testing, associations with age and respiratory rate remained significantly associated with mortality (Figure 2).

### D-dimer ≥1,000 μg/L

We chose not to include D-dimer (<1000 μg/L and ≥1,000 μg/L) in multivariable models for admission to AICU, mechanical ventilation, and mortality due to the degree of incomplete data for D-dimer, such that restricting analyses to patients with complete data would have resulted in a significant reduction in cases and thus statistical power (Supplementary figure 1). In the Fisher’s exact tests, unadjusted for potential confounders D-dimer ≥ 1,000 μg/L was associated with approximately 9-fold greater odds of AICU admission (OR = 9.041, 95% CI: 5.499-15.449) and 8-fold greater odds of mechanical ventilation (OR = 8.390, 95% CI: 4.553-16.504), but was not associated with mortality (OR = 1.341, 95% CI: 0.948-1.893). 2−2 contingency tables used to compute ORs can be found in Supplementary Table 3.

We observed increased odds of an elevated recorded D-dimer measurement (≥1,000 μg/L) in patients with higher serum potassium levels (OR = 1.615 per 1 mmol/L, 95% CI: 1.091-2.464), higher CRP levels (OR = 1.007 per 1mg/L, 95% CI: 1.005-1.010), higher numbers of white blood cell numbers (OR = 1.093 per 10^9^/L, 95% CI: 1.035-1.162), and other/unspecified ethnic background (OR = 1.811, 95% CI: 1.032-3.209). Higher levels of haemoglobin were associated with lower odds of elevated D-dimer (OR = 0.982 per 1 g/L, 95% CI: 0.970-0.993)

In the multivariable model developed in 485 patients with 368 cases of at least one elevated D-dimer measurement, results indicated that older age (OR = 1.023 per 1 year, 95% CI: 1.004-1.043), elevated CRP (OR = 1.004 per 1 mg/L, 95% CI: 1.002-1.006), and higher body temperature (OR = 0.723 per 1°C, 95% CI: 0.541-0.958) had small incremental associations with an elevated D-dimer measurement. No significant associations with elevated D-dimer were observed with sex, and all other associations were attenuated by multivariable adjustment. Following the adjustment for multiple hypothesis testing, only CRP remained significantly associated with elevated D-dimer (Figure 2).

## Discussion

This retrospective analysis included 2756 patients, of whom 1148 were confirmed positive and 1608 were confirmed negative for COVID-19 at admission to the hospital emergency department. We have identified several factors associated with a symptomatic presentation of COVID-19, as well as likelihood of progression from mild to severe illness defined by AICU admission, mechanical ventilation, in-hospital mortality, and at least one recorded D-dimer measurement of ≥1,000 μg/L.

Consistent with other recently published observational studies, we found that patients with COVID-19 were more likely to be older, male, and to present with one or more comorbidities previously diagnosed in the Trust, when compared to other patients admitted to the hospital without COVID-19 (15). Specifically, we found that history of hypertension and type-II diabetes were more prevalent among COVID-19 patients that had a record in our data of any medical diagnosis previously attributed in the Trust (15). Indeed, hypertension and type-II diabetes are widely reported to be the most common among patients with COVID-19 and are believed to increase the risk of severe complications including ARDS and multi-organ failure (16, 17).

When comparing laboratory measurements recorded at the time of patient admission to the hospital’s ED, we observed several haematological biomarkers of importance that were significantly increased or reduced among patients with COVID-19. Notably, we found patients with COVID-19 had higher levels of seven biomarkers associated with inflammation, organ function and tissue damage (lung, heart, kidney, and liver), including CRP, LDH, ferritin, ALT, creatine kinase, and creatinine. These observations match those of other studies (18, 19). Further, we observed systematically lower absolute numbers of lymphocytes, neutrophils, and monocytes in patients with COVID-19.

When analysing the risk factors associated with presentation with symptomatic COVID-19, we identified nine parameters positively associated with COVID-19 positive status: older age, male sex, self-reported Asian or un-specified ethnicity, increased levels of CRP, higher body temperature, reduced numbers of white blood cells, and lower respiratory rate and systolic blood pressure. Predictive models incorporating these risk factors may therefore be valuable in supporting patient triage and prioritisation of treatment administration. Our results align with those of previous studies, which have shown associations between older age, male sex, unmediated inflammatory response to infection, and lymphopenia with increased risks of AICU admission for severe complications (18, 20, 21).

In our multivariable analysis, we explored the existence of associations between key risk factors and severe COVID-19 disease progression, here measured as admission to AICU, mechanical ventilation, elevated D-dimer measurements, and death. Our results demonstrate that higher levels of CRP increased the risk of all outcomes, indicating that virally driven hyperinflammation may play a key role in driving inferior prognoses (6, 21, 22). Moreover, D-dimer ≥1,000 μg/L was associated with increased risk of AICU admission and mechanical ventilation. Elevated D-dimer can be indicative of increased risk of venous thromboembolism, as well as of indicative of other conditions such as inflammation, sepsis, and infection, and has been more recently associated with poor outcomes in COVID-19 patients (23, 24). However, its lack of association with mortality warrants further investigation.

Analyses also showed that older age and increased levels of sodium are inversely associated with both admission to AICU and mechanical ventilation. Although these findings have not yet been independently corroborated by other studies in this area, researchers have recently postulated that high dietary sodium may downregulate expression of angiotensin-converting enzyme 2 (ACE-2) receptor, by which SARS-CoV is known to infect host cells (25). This hypothesis may help to explain the inverse associations seen in our study, but further investigations are required, particularly in light of the fact that increased risk of mortality was observed in patients with higher serum levels of sodium. Interestingly, while older age was protective for admission to the AICU and mechanical ventilation, it was also associated with greater risk of mortality. Risk of mortality was further elevated in males, those of Asian ethnic background, and those with a higher respiratory rate at admission to the hospital.

Our work was able to shed new light on the clinical profile of COVID-19 patients and factors associated with inferior prognoses. However, the degree of incomplete data for patients’ medical and treatment history, as well as for laboratory measurements and vital sign observations recorded at the time of patients’ admission to the hospital, may have introduced bias into our analyses when comparing only those patients with available data. Many patients will have received diagnoses and medications in a primary care setting or in other hospital Trusts, which are sources of data that were not available for use in this study. In addition, the two hospitals used in this study have had a staggered deployment of their EHR such that one site is understood to not record certain data related to mechanical ventilation. From clinical feedback the total number of mechanically ventilated patients was 135, however only 71 were visible in our data. Further work will be needed to address these aspects.

Moreover, as expected in a real-world evidence setting, particularly in the midst of a pandemic, coverage of laboratory measurements and vital signs were more extensive in COVID-19 patients (25.8% to 85.1%) than in patients without COVID-19 (5.9% to 27.7%). Hence, it is possible that patients without COVID-19 for whom these measurements were recorded had worse health states or inferior prognoses, thus resulting in more conservative estimates of associations in our analyses when comparing differences in characteristics with patients with and without COVID-19. Similar concerns extend to our analysis of AICU admission, mechanical ventilation, and elevated D-dimer. In restricting analyses to patients with complete data, we may have included patients systemically different from the remaining population, as well as constrained analyses by reducing statistical power. However, many associations were biologically plausible and remained statistically significant following adjustment of P-value significance thresholds with the Holm-Bonferroni correction method, which adds weight to the validity of our findings. We must also acknowledge the likelihood of residual confounding in this study, as there were many risk factors, including but not limited to smoking, body-mass index, and medical history, that we were not able to adjust for in analyses.

This study includes a larger sample size compared to other work in this area and includes a comprehensive assessment of associations of an extensive panel of clinical measurements readily measured at patient admission with COVID-19 status and clinical deterioration. Our study provides further evidence and support to existing literature of key risk factors for one or more severe outcomes in patients with COVID-19.

## Data Availability

The data are not publicly available

## Acknowledgments

This work uses data provided by patients and collected by the NHS as part of their care and support. We believe using patient data is vital to improve health and care for everyone. The authors would, thus, like to thank all those involved for their contribution.

## Ethics statement

The data were extracted, anonymised, and supplied by the Trust in accordance with internal information governance review, NHS Trust information governance approval, and General Data Protection Regulation (GDPR) procedures outlined under the Strategic Research Agreement (SRA) and relative Data Sharing Agreements (DSAs) signed by the Trust and Sensyne Health plc on 25th July 2018.

